# Vaccination strategies impact the probability of outbreak extinction: a case study of COVID-19 transmission

**DOI:** 10.1101/2022.07.23.22277952

**Authors:** Natcha C. Jitsuk, Sudarat Chadsuthi, Charin Modchang

## Abstract

Mass vaccination has been one of the effective control measures for mitigating infectious disease transmission. Several vaccination strategies have been introduced throughout history to control infections and terminate the outbreak. Here, we employed the coronavirus disease 2019 (COVID-19) transmission as a case study and constructed a stochastic age-structured compartmental model to investigate the effectiveness of different vaccination strategies. We estimated the outbreak extinction probability under different vaccination scenarios in homogeneous and heterogeneous populations. We found that population heterogeneity could enhance the likelihood of outbreak extinction at various vaccine coverage. In addition, prioritizing vaccines for people with higher infection risk could maximize the outbreak extinction probability and reduce more infections. In contrast, allocating vaccines to individuals with higher mortality risk provides better results in reducing deaths. We also found that as the vaccine effectiveness wane over time, a booster dose of vaccine could significantly enhance the extinction probability and mitigate disease transmission.

## 1. Introduction

Infectious diseases have continuously afflicted humankind throughout history [1–6]. Today, humanity faces the latest infectious disease since late 2019, the coronavirus disease 2019 (COVID-19) [1, 7]. After the World Health Organization (WHO) declared the novel infectious disease had emerged and it trended to spread across expansive geographical areas, many non-pharmaceutical interventions were implemented to prevent and control the spread of COVID-19 [8, 9]. Even though the non-pharmaceutical control measures could reduce the risk of infection and transmission, massive vaccination always remains the foreseeable hope for returning to old normal behavior and ending the COVID-19 pandemic. The ultimate goal of disease control is to eradicate infections, and mass vaccination could, in principle, terminate the disease transmission [10–13].

In early 2021, the COVID-19 vaccines were successfully developed and started to roll out to populations worldwide [14]. Some governments faced, however, several hurdles in prioritizing vaccine allocation at the first phase of vaccine dissemination. Some countries granted the frontline healthcare workers assessing vaccines as the first tier because they could protect the high-risk population. Some nations decided to disseminate the vaccine to those at the highest risk of dying. The vaccination plans are, therefore, developed to encounter the challenge of distributing several million vaccine doses.

To assess the optimal vaccination plan, mathematical models are an effective tool for predicting the suitable approach for several goals, such as maximizing the reduction in mortality, infection, or hospitalization and achieving herd immunity [15–18]. Bubar K. M. *et al*. recently proposed a strategy to vaccinate prioritized by age-stratification [15]. Their results indicated that the vaccines prioritized for 20-49 years old could effectively minimize the incident cases, and the number of deaths decreases if the vaccines are prioritized for elderlies of age more than 60-year-old [15]. Markovič R. *et al.* found that prioritizing vaccines for seniors and other high-risk groups is beneficial only if there is no vaccine limitation. However, it would be better if healthy individuals were the first tier for getting vaccinated under the limited vaccine accessibility scenario [18].

After massive vaccination was, however, implemented to control the COVID-19 transmission, several studies revealed that vaccine effectiveness wanes over time [19–22]. Moreover, new SARS-CoV-2 variants could escape the immune and surge infections. It has been shown that the virus could mutate and elude vaccine-induced or natural immunity. Consequently, it might affect disease transmission, the number of deaths, as well as the chance of outbreak going extinct and, therefore, could drive the COVID-19 to re-emerge in the future.

In this study, we aimed to investigate the impact of various vaccination strategies on the probability of outbreak extinction by employing the COVID-19 transmission as a case study. Besides, we examined the impact of vaccination strategies on infections and deaths. We also examined the robustness of the vaccination plans by varying the contact matrices and the population structures. Furthermore, we studied the effect of the waning vaccine effectiveness on change in the probability of disease extinction and disease transmission dynamics.

## 2. Materials and Methods

### 2.1 Model Structure

We applied the extended SEIR model to illustrate the COVID-19 transmission. From the characteristic of COVID-19, the individuals in the system are classified into thirteen compartments depending on their health status (**Figure 1(A)**). The compartments *S*, *L*, *I*, *A*, *R*, and *D* are for unvaccinated populations in the susceptible, latent, symptomatic infectious, asymptomatic infectious, recovered, and dead populations, respectively. The remaining compartments contain fully vaccinated populations. *S*_P_, *L*_P_, *I*_P_, *A*_P_, *R*_P_, and *D*_P_ represent the imperfectly immunized symptomatic infectious, asymptomatic infectious, recovered, and dead individuals, respectively. The individuals in these compartments could be infected by others or transmit the disease to others. *S*_V_ refers to the vaccinated people who get perfect immunity against infection; therefore, they are not infected by other infectious individuals.

**Figure 1:**
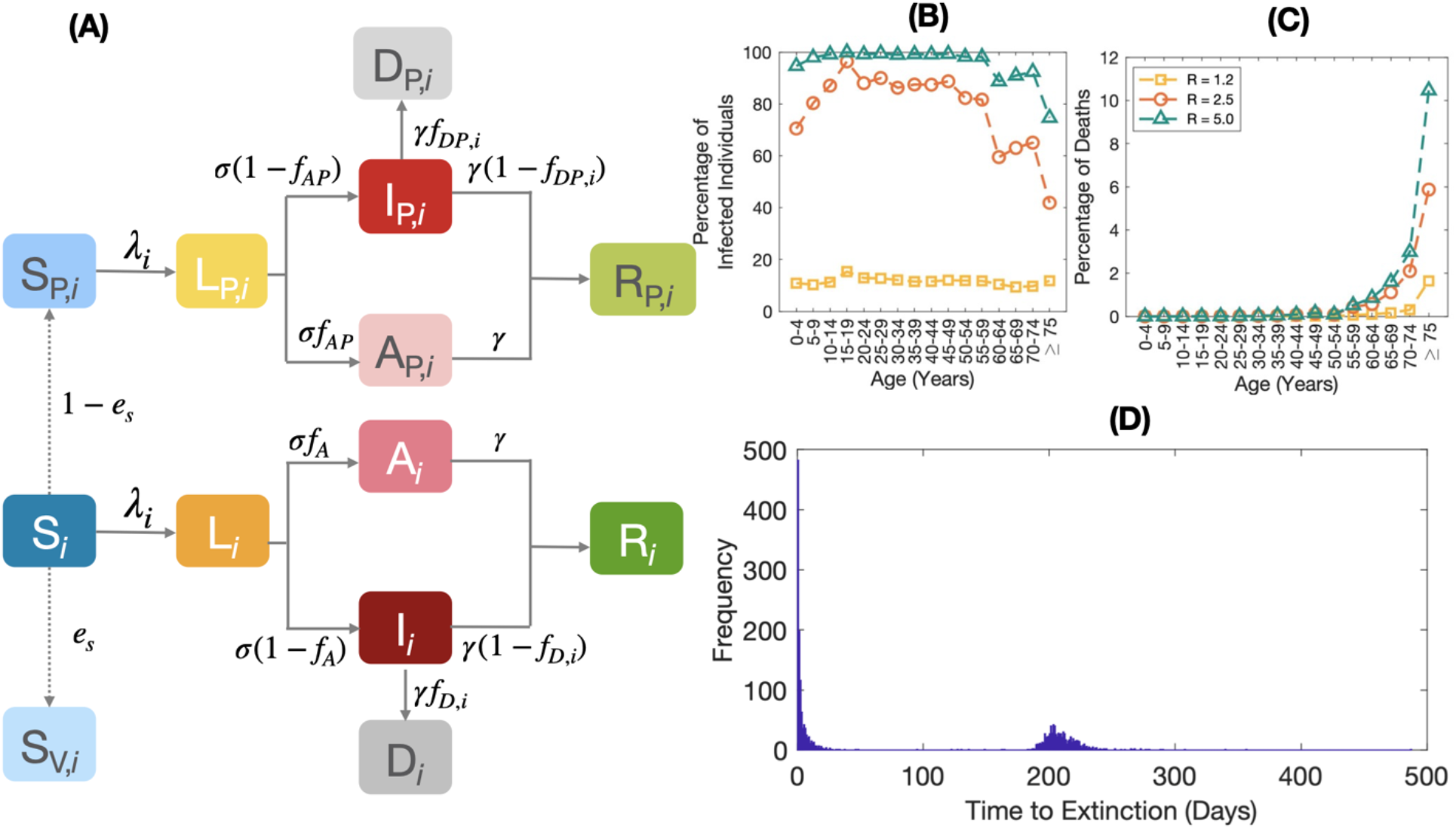
**(A)** The disease transmission with vaccination constraint model. The solid lines show the transition between two compartments. **(B)** and **(C)** The infection risk and the mortality risk at a different value of *R*. **(D)** The frequency of extinction obtained from 2,000 model realizations, under the conditions of *R* = 2.5 and 10% of vaccine coverage.

The age structure is incorporated into the model by dividing the population in each epidemiological compartment into 16 age groups: 0-4, 5-9, 10-14, 15-19, 20-24, 25-29, 30-34, 35-39, 40-44, 45-49, 50-54, 55-59, 60-64, 65-69, 70-74 and above 74 years old. People in each age group contact people in other groups by the different contact rates, leading to the heterogeneity in the system. In this study, the data on the age structure of populations and the contact matrices are obtained from [23, 24]. A *who acquires infection from whom* (WAIFM) matrix, ***M*** = [*M_ij_*], is employed to describe the number of contacts of the populations from the *i*th age group to the individuals in the *j*th age group. The transmission matrix, ***β*** = [*β_ij_*], is the product of the contact matrix ***M*** and a scaling factor *κ*,

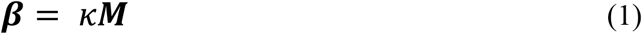

The next-generation matrix, ***N***, is given by

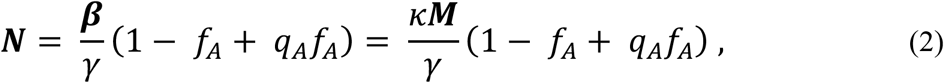

Where *q*_A_ is a parameter representing the relative infectiousness of asymptomatic infectious individuals. *f*_A_ is the fraction that unvaccinated latently infected population becomes asymptomatic. *γ* is the recovery rate.

The basic reproductive number is defined as the maximum eigenvalue of the next generation matrix [25].

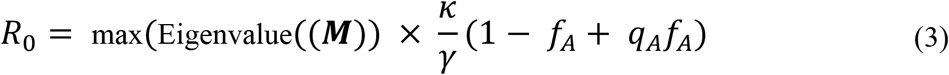

Consequently, and the transmission matrix is calculated from

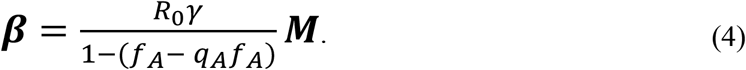

The population in *S* and *S*_P_ compartments can get infected from the populations in *I*_P_, *A*_P_, *I*, and *A* compartments. After getting infections, the individuals in *S* and *S*_P_ compartments become the population in *L* and *L*_P_ compartments, respectively, with a transition rate known as the force of infection (*λ*). The force of infection of individuals in age group *i* is given by

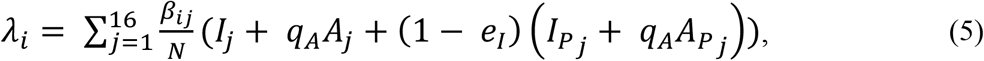

where *N* is the total population, *q*_A_ is a parameter representing the relative infectiousness of asymptomatic infectious individuals, *e*_I_ is the vaccine effectiveness against transmission. The individuals in *L* and *L*_P_ compartments become the asymptomatic infectious populations in *A* and *A*_P_ compartments with the transition rate *σ* which equals the reciprocal of the latent period. *f*_AP_ is the proportion that the population in the *L*_P_ compartment becomes asymptomatic and is calculated from (see Supplementary Information for more details)

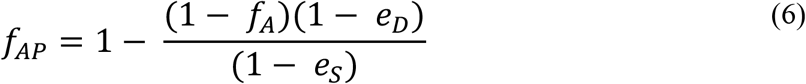

*e*_D_ and *e*_S_ are the vaccine effectiveness against symptomatic disease and infection, respectively [26–29]. All asymptomatic infectious individuals become recovered with the recovery rate, *γ*.

The population in *L* and *L*_P_ compartments becomes the symptomatic infectious populations in *I* and *I*_P_ compartments. The symptomatic populations die with transition rate *γ*. *f_D,i_* and *f_DP,i_* are the proportions which unvaccinated and the vaccinated infected populations in age group *i* die due to the infection, respectively, and was estimated from (see Supplementary Information for more details)

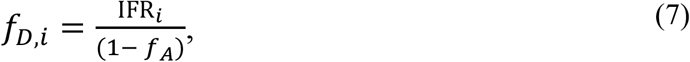

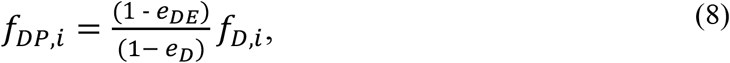

where IFR*_i_* is the infection fatality ratio of infected individuals in age group *i* obtained from [15, 30]. *e_DE_* is the vaccine effectiveness against death.

The values of parameters used in the model are summarized in **Table 1.**

**Table 1:** The parameters and their default values used in the model.

| Parameters | Definition | Value | References |
| --- | --- | --- | --- |
| $\beta$ | Transmission matrix | Eq. (4) | - |
| $\sigma$ | Transition rate of individuals in the latent compartment to the infectious compartment | 1/3 day<sup>-1</sup> | [31] |
| $\gamma$ | Recovery rate | 1/2.5 day<sup>-1</sup> | [31] |
| $q_A$ | Relative infectiousness of asymptomatic infectious individuals | 0.58 | [32-34] |
| $f_{AP}$ | Proportion of asymptomatic infections for vaccinated individuals | Eq. (3) | - |
| $f_A$ | Proportion of asymptomatic infections for unvaccinated individuals | 0.40 | [32-35] |
| $f_{DP,i}$ | Proportion of symptomatic breakthrough-infected individuals in age group $i$ who eventually die | Eq. (4) | - |
| $f_{D,i}$ | Proportion of symptomatic infected individuals in age group $i$ who eventually die | Eq. (5) | - |
| $e_I$ | Vaccine effectiveness against transmission | 0.032 | [36] |
| $e_S$ | Vaccine effectiveness against infection | 0.55 | [37] |
| $e_D$ | Vaccine effectiveness against disease | 0.49 | [38] |
| $e_{DE}$ | Vaccine effectiveness against death | 0.95 | [39] |

### 2.2 Vaccination Strategies

In this study, five vaccination strategies: i.e., 1) uniform, 2) infection-risk priority, 3) infection-risk weighted, 4) mortality-risk priority, and 5) mortality-risk weighted, are proposed and examined their effect on the outbreak extinction probability, the number of cases, and the number of deaths. The details of these vaccination strategies are as follows:

- The uniform vaccination strategy: individuals in all age groups have an equal chance to get vaccinated.
- The infection-risk priority strategy: vaccines are prioritized for all individuals in the group with a higher risk of infection, while people in other groups with a lower infection risk get vaccinated later until all allocated vaccines are used up.
- The infection-risk weighted vaccine allocation: vaccines are shared with every age group with different portions, weighted by the risk of infection. In this sense, the group with higher risk will get more vaccine doses than those at lower risk.
- The mortality-risk priority and mortality-risk weighted vaccination strategies: the vaccine is rolled out similar to the infection-risk priority and infection-risk weighted vaccination strategies, respectively, but the vaccine is prioritized and weighted by the mortality risk.

To identify the infection risk and the mortality risk of individuals in each age group, we estimated the risk of infection and mortality as the number of infected individuals and the number of deaths in the age group divided by the population of that age group, respectively. The number of cumulative infected individuals and the number of cumulative deaths are estimated from the disease transmission model under 0% vaccine coverage condition. The infectious disease spread continuously until the cumulative cases reached equilibrium. Then, we get the number of infected individuals and the number of deaths in each age group to estimate the infection risk and the mortality risk, as shown in **Figures 1(B) - 1(C)**.

### 2.3 Simulation Details

Here, the effectiveness of vaccination strategies is assessed using the probability of outbreak extinction, the cumulative cases, and the cumulative deaths. To measure the extinction probability, the population is initially set in *S*, *S*_P_, and *S*_V_ compartments. The unvaccinated population is in the S compartment. Some vaccinated individuals are in the *S*_V_ and *S*_P_ compartments with the proportion of *e*_S_ and 1-*e*_S_ of the total vaccinated population, respectively. At the initial time, there is only one symptomatic or asymptomatic infectious individual in the system. In this study, we employ the tau-leaping method to simulate disease transmission. When there are no infectious individuals in the system, the simulation stops, and the extinction time is recorded. The simulation algorithm is summarized in **Figure S1**.

After the extinction time of each model realization is recorded, we plot the frequency of the extinction time, as shown in **Figure 1(D)**. As can be seen, the extinction times can be separated into two groups. The left group is for the simulations in which the disease transmission becomes extinct. On the other hand, the right group is for the simulations in which the disease could spread continuously and infect a substantial number of individuals. The probability of extinction is computed from the number of simulations on the left group divided by the total number of simulations. All figures and calculations are generated by using MATLAB software (version R2020b; The MathWorks, Inc).

### 2.4 Waning Vaccine Effectiveness

To examine the impact of waning vaccine effectiveness on the extinction probability, relative case, and relative death, we employ the method for investigating the extinction probability as abovementioned. The vaccine effectiveness against the Omicron variant waned each month after getting the full protection of the vaccine regimen was set as the initial condition, see **Table S2**.

## 3. Results

### 3.1 Effect of population heterogeneity

We first validated the method for investigating the extinction probability by comparing the simulation results of the extinction probability in a homogeneous system in which the contact patterns between age groups are uniform to those obtained from the existing theory where the extinction probability (*P_ext_*) is given by [40]

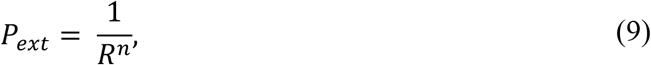

where *R* is the effective reproductive number and *n* is the initial number of infectious individuals.

As shown in **Figure 2(A)**, the extinction probabilities at different values of reproductive number, *R*, are plotted against vaccine coverage. The marked lines represent the computational results, and the dashed lines depict the theoretical results. The two corresponding lines are well-traced regarding different values of *R* and all percentages of the vaccinated population, showing the rigorous agreements between the two approaches. These comparable results validate the methods and the criteria for numerically estimating the probability of outbreak extinction.

**Figure 2:**
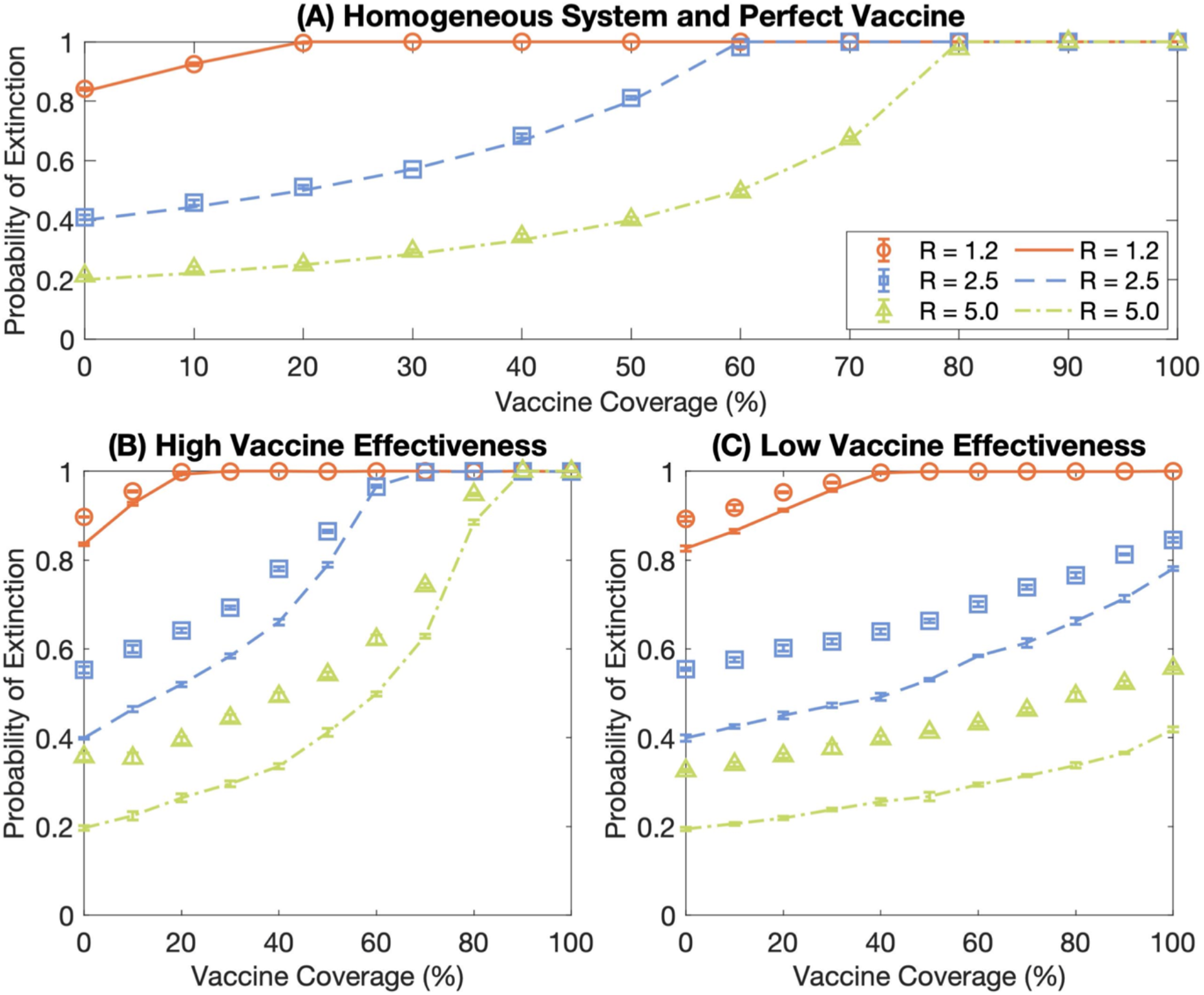
The comparison of the probability of extinction in the heterogeneous and homogeneous systems. The orange-circle, blue-square, and green-triangle marks represent the extinction probabilities of the heterogeneous system at *R* = 1.2, 2.5, and 5.0, respectively. The orange-solid, blue-dashed, and green-dash-dotted lines show the probability of extinction of the homogeneous system at *R* = 1.2, 2.5, and 5.0, respectively. Error bars indicate the standard error of the mean. **(A)** displays the comparison of the probability of extinction between the simulation results and the theoretical results under the perfect vaccine and homogeneous population conditions. **(B)** illustrates the extinction probabilities of the high vaccine effectiveness scenarios. **(C)** presents the extinction probabilities of the low vaccine effectiveness scenarios.

We then examined the impact of the population heterogeneity on the extinction probability under imperfect vaccine scenarios. The extinction probability in the heterogeneous system compared with those in the homogeneous system is presented in **Figures 2(B) - 2(C)**. All marks represent the probabilities in the heterogeneous system, and the lines illustrate such probabilities in the homogeneous system. **Figures 2(B) - 2(C)** display the extinction probabilities under the high vaccine effectiveness (against the Alpha variant (B.1.1.7), see **Table S1**) and the low vaccine effectiveness (against the Omicron variant (B.1.1.529), see **Table 1**), respectively. We found that the extinction probabilities at all vaccine coverages in the heterogeneous system are consistently higher than those in the homogeneous system.

### 3.2 Impact of vaccination strategies

We investigated the effect of different vaccination strategies on the probability of outbreak extinction, the number of cases, and the number of deaths. Five vaccination strategies were investigated in this study: i.e., uniform, infection-risk priority, infection-risked weighted, mortality-risked priority, and mortality-risked weighted vaccination strategies. **Figure 3(A)** illustrates the probability of outbreak extinction, the number of cases per 100,000 population, and the number of deaths per 100,000 population of each vaccination strategy under a low vaccine effectiveness scenario, respectively. We found that various vaccine distribution strategies impact the probability of extinction significantly. As shown in **Figures 3(A1)** – **3(A2)**, the infection-risked priority strategy could maximize the extinction probability compared to other vaccination plans with the same vaccine coverage. Correspondingly, this strategy could remarkably reduce the cumulative cases. For curbing the number of deaths, both mortality-risk priority and mortality-risk weighted vaccine allocation strategies show good results because individuals in the high mortality risk group are the first tier for getting the vaccine (**Figure 3(A3)**). It, therefore, influences the reduction in deaths efficiently. However, the results depict the vaccine coverage threshold for reducing the number of deaths. It indicates that the number of deaths becomes minimum even if the vaccine coverage is higher than the threshold. It is due to all high mortality-risk populations getting vaccinated.

**Figure 3:**
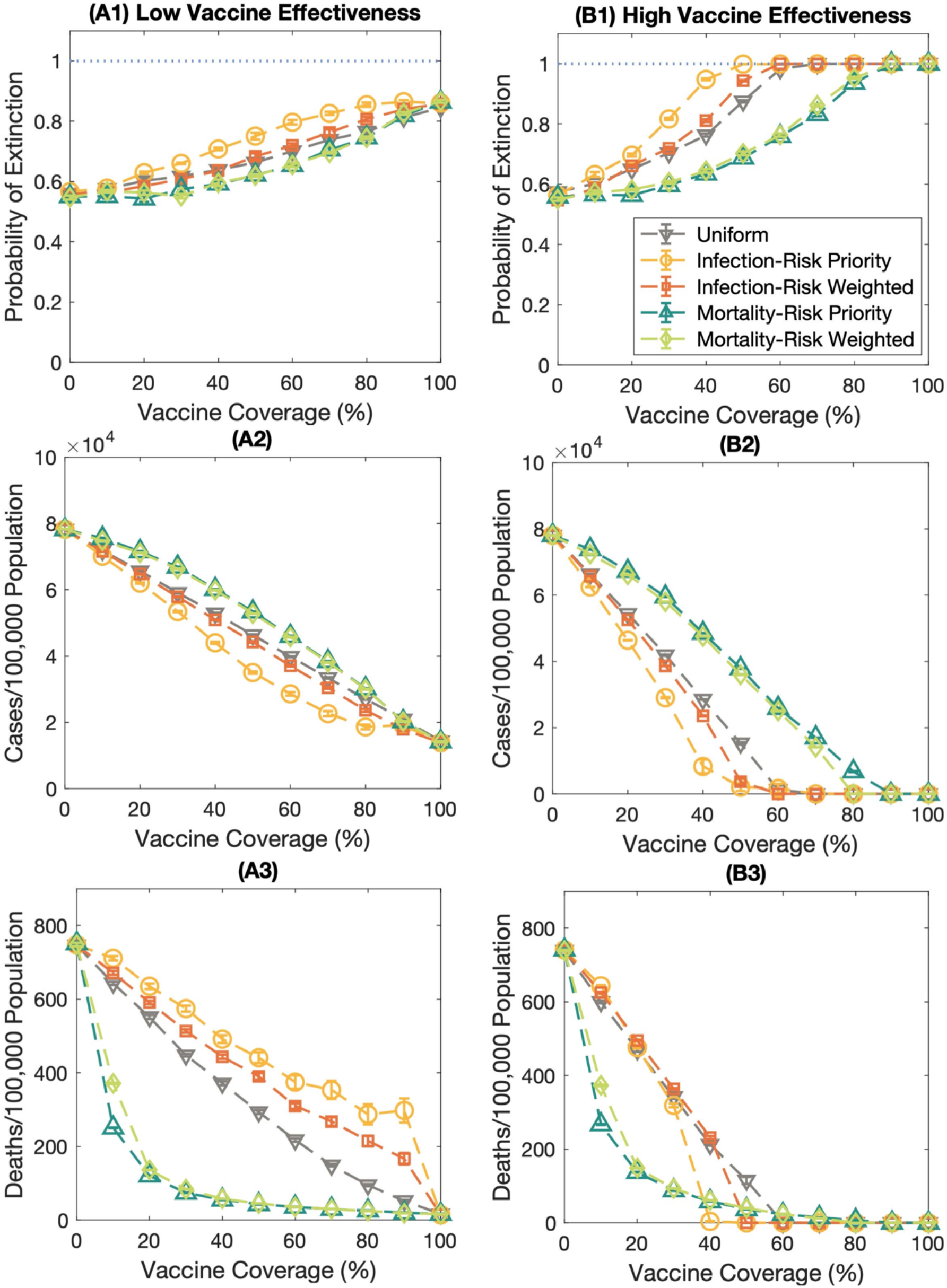
The impact of various vaccination strategies on the probability of extinction, the number of cases per 100,000 population, and the number of deaths per 100,000 population. The dashed lines with the downward-pointing triangle, the circle, the square, and the upward-pointing triangle, and the diamond marks represent the probability of extinction under the uniform, infection-risk priority, infection-risk weighted, mortality-risk priority, and mortality-risk weighted vaccination strategies, respectively. Error bars represent the standard error. **(A)** shows the results under the low vaccine effectiveness scenario. **(B)** display the results under the high vaccine effectiveness condition.

For the high vaccine effectiveness scenario, the infection-risk priority vaccination plan still shows outstanding performance to maximize the extinction probability with the minimum vaccine doses and curb the cumulative cases, see **Figures 3(B1)** – **3(B2)**. Moreover, the infection-risk priority vaccination strategy could relieve the number of deaths when the vaccine coverage is high enough. For low vaccine coverage, either mortality-risk priority or mortality-risk weighted strategies are the appropriate solutions to mitigate the mortality rate, as indicated in **Figure 3(B3)**.

When disease transmission is high, the vaccination plan might not affect the extinction probability and the infection rate because the disease could be transmitted fast. Population in each age band could get infections with a high chance as same as others (**Figures S2(A) – S2(B)**). Although various vaccine allocation strategies do not impact the extinction probability and the cumulative cases much, the vaccination strategies still influence the reduction in the number of deaths. We disclosed that both mortality-risk priority and mortality-risk weighted vaccination strategies could mitigate the rate of dying even if the disease spreads fast, **Figure S2(C)**.

### 3.3 Effect of population structure and contact pattern

From the previous section, the results reveal that the infection-risk priority vaccination strategy might be a suitable strategy leading the outbreak to go extinct with the minimum vaccine coverage compared to other vaccination strategies. This strategy is also the optimal option for minimizing the number of cumulative cases. However, the infection-risk priority vaccination strategy might not be a suitable alternative for mitigating the mortality rate. The results indicate that the mortality-risk priority and the mortality-risk weighted vaccination strategies are more appropriate for reducing the rate of dying.

Mass vaccination is one of the effective ways to constrain infectious disease transmission. Various vaccination plans were proposed to allocate the vaccine in multiple nations. Here, we examined whether the infection-risk priority vaccination strategy is a suitable plan to alleviate the number of cases and whether both mortality-risk priority and mortality-risk weighted vaccination strategies are the proper ones to mitigate the deaths in various territories. Four countries with different population structures and contact patterns, namely, Ethiopia, India, Thailand, and the United Kingdom, were employed as examples of this study.

The results uncover that the infection-risk priority vaccination strategy is still the effective vaccination plan for maximizing the extinction probabilities for several contact patterns (**Figure 4** Moreover, the infection-risk priority vaccination strategy could relieve the infection rate (**Figure S3)**. To reduce the mortality rate, the mortality-risk priority and the mortality-risk weighted vaccination strategies still achieve better performance than other vaccination strategies (**Figure 5)**. However, there is a threshold for vaccine coverage in which the number of deaths per 100,000 population becomes minimum even if the vaccine coverage is higher. These results are consistent for all contact patterns and population structures, but the consequence of the uniform and the infection-risk weighted vaccine allocation plans are dissimilar.

**Figure 4:**
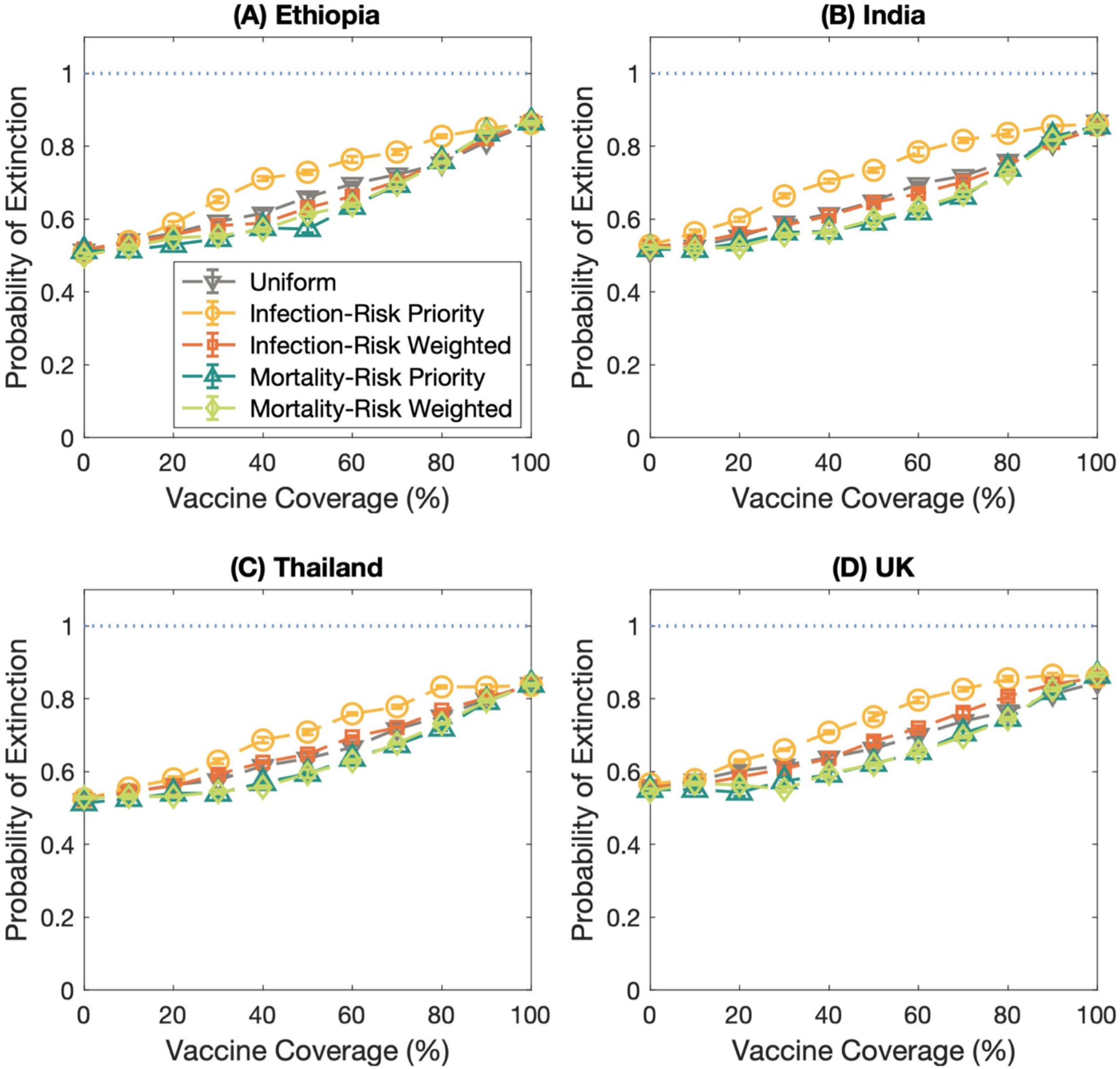
The probability of extinction in different countries; **(A)** Ethiopia, **(B)** India, **(C)** Thailand, and **(D)** the United Kingdom. The dashed lines with the downward-pointing triangle, the circle, the square, and the upward-pointing triangle, and the diamond marks represent the probability of extinction under the uniform, infection-risk priority, infection-risk weighted, mortality-risk priority, and mortality-risk weighted vaccination strategies, respectively. Error bars show the standard error.

**Figure 5:**
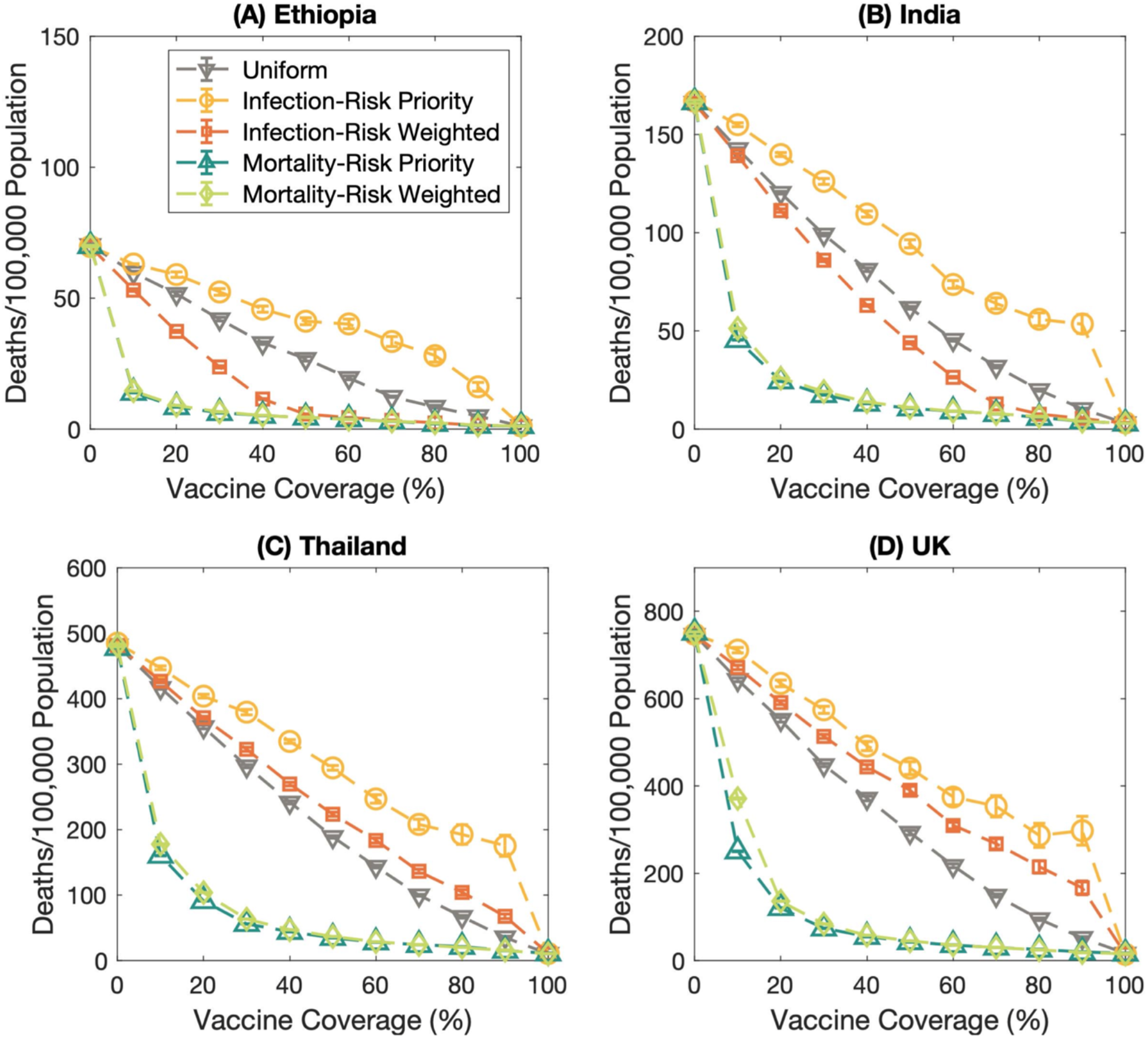
The number of deaths per 100,000 population in various countries; **(A)** Ethiopia, **(B)** India, **(C)** Thailand, and **(D)** the United Kingdom. The dashed lines with the downward-pointing triangle, the circle, the square, and the upward-pointing triangle, and the diamond marks represent the number of deaths per 100,000 population of the uniform, infection-risk priority, infection-risk weighted, mortality-risk priority, and mortality-risk weighted vaccination strategies, respectively. Error bars show the standard error.

### 3.4 Effect of waning vaccine effectiveness

Several studies revealed that the COVID-19 immunity induced by the vaccine is wanned over time. Here, we studied the impact of waning vaccine effectiveness on the probability of outbreak extinction, the relative case, and the relative death. **Figures 5 – 7** represent the effect of waning vaccine effectiveness against the omicron variant on the probability of outbreak extinction, the relative case, and the relative death at various times since getting full protection from the full doses of the COVID-19 vaccine, respectively. **Figures 5(A) – 5(D)** illustrate the extinction probabilities under the varied vaccine coverage: 25%, 50%, 75%, and 90%, respectively. As expected, the waning vaccine effectiveness influences the probability of extinction. The results reveal that the probability of extinction is the greatest in the first month after getting full protection from the complete regimen and after the booster dose. At 2 and 3 months, the probability of extinction almost equals the extinction probability of 0% vaccine coverage condition, even though the vaccine coverage is high. Moreover, the results demonstrate that the different vaccination strategies do not affect the extinction probability when the vaccine effectiveness extremely wanes.

**Figure 6:**
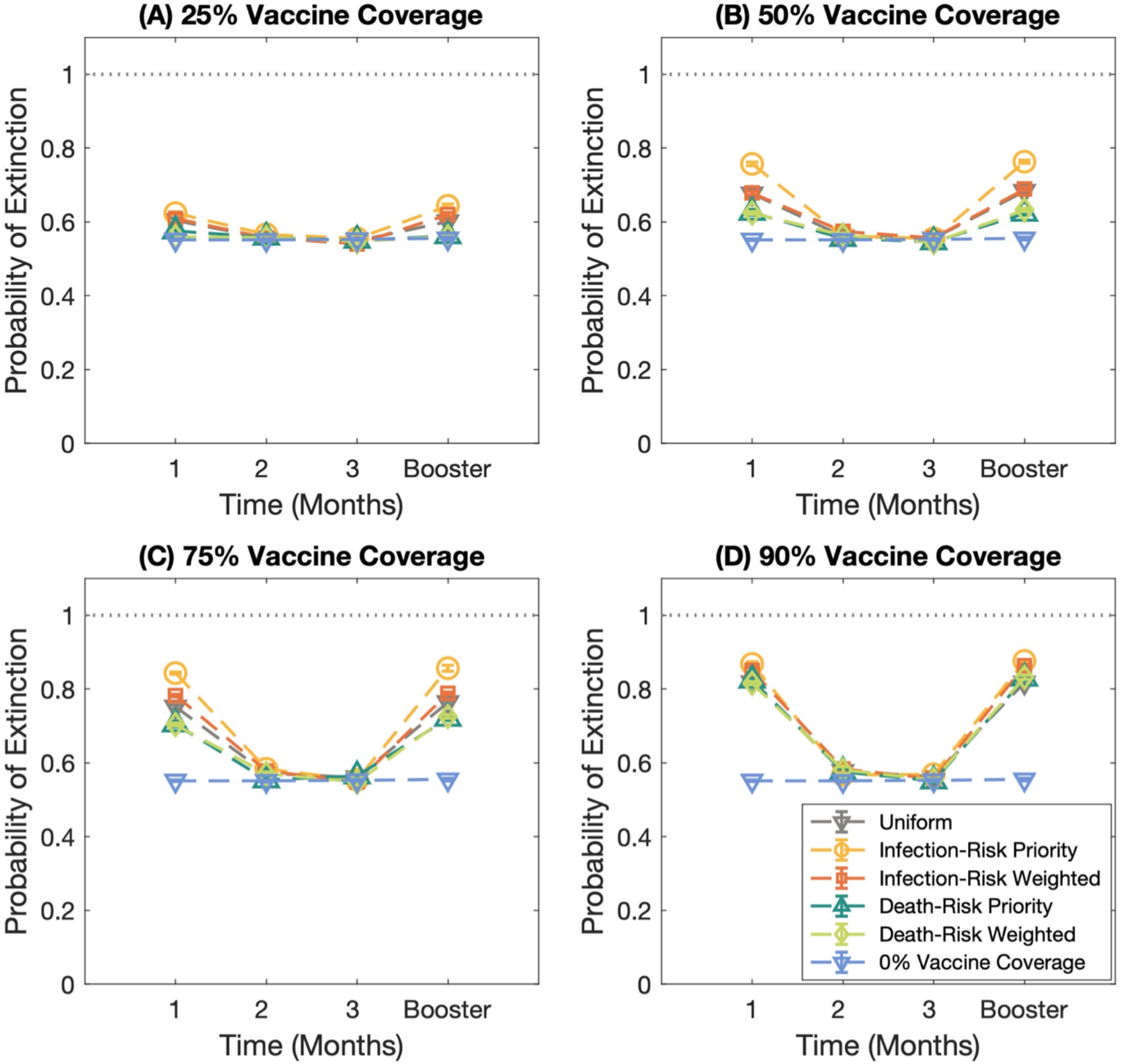
The effect of waning vaccine effectiveness against the Omicron variant (B.1.1.529) on the probability of outbreak extinction. **(A) – (D)** display the extinction probabilities under the different vaccine coverages; 25%, 50%, 75%, and 90% of the total population, respectively. Each line represents the extinction probability of different vaccination strategies. The horizontal dash line with the downward-pointing triangle marks displays the probability of extinction under 0% vaccine coverage.

**Figure 7:**
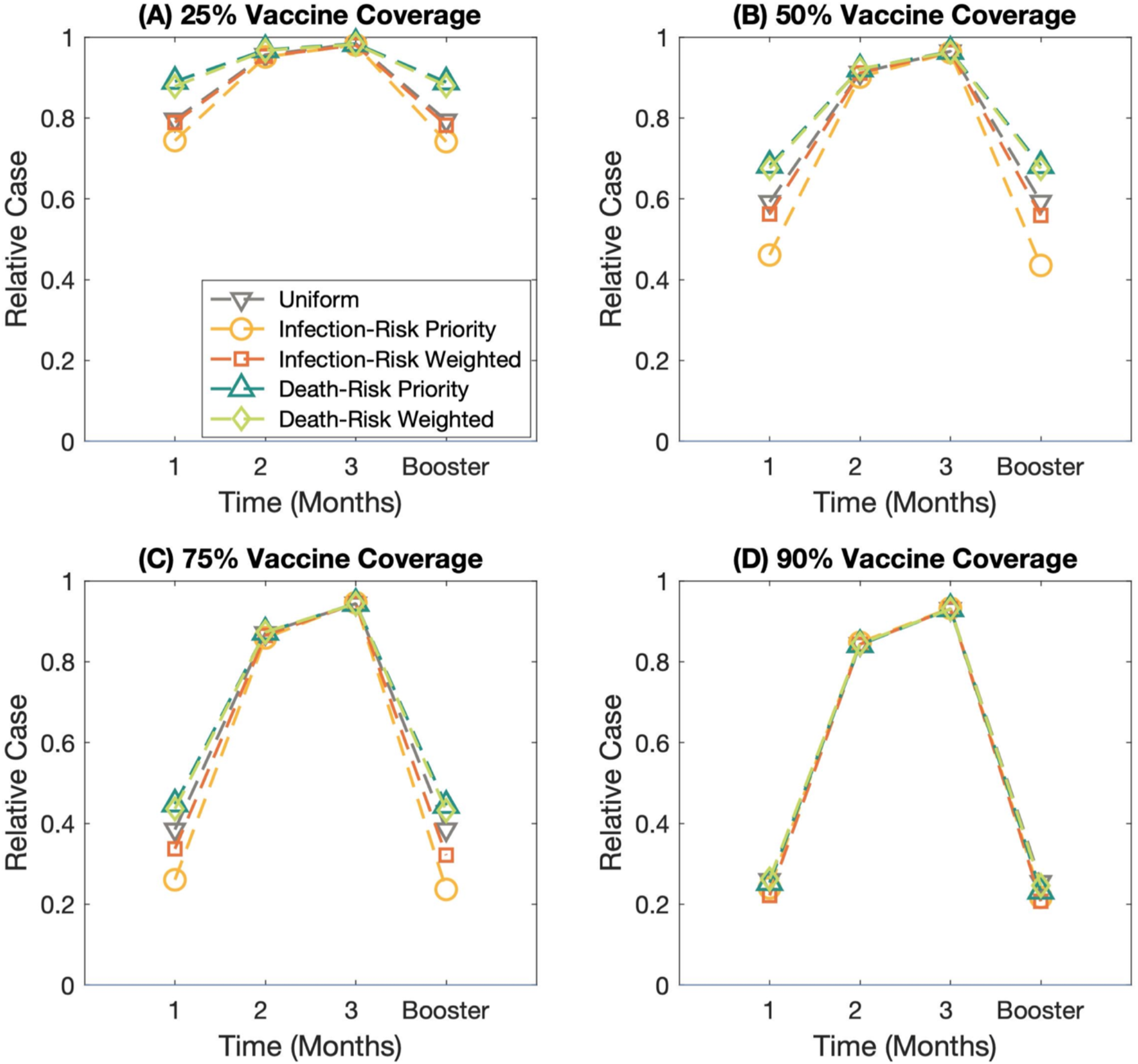
The effect of waning vaccine effectiveness against the Omicron variant (B.1.1.529) on the relative case. **(A) – (D)** display the extinction probabilities under the different vaccine coverages; 25%, 50%, 75%, and 90% of the total population, respectively. Each line represents the extinction probability of different vaccination strategies.

**Figure 8:**
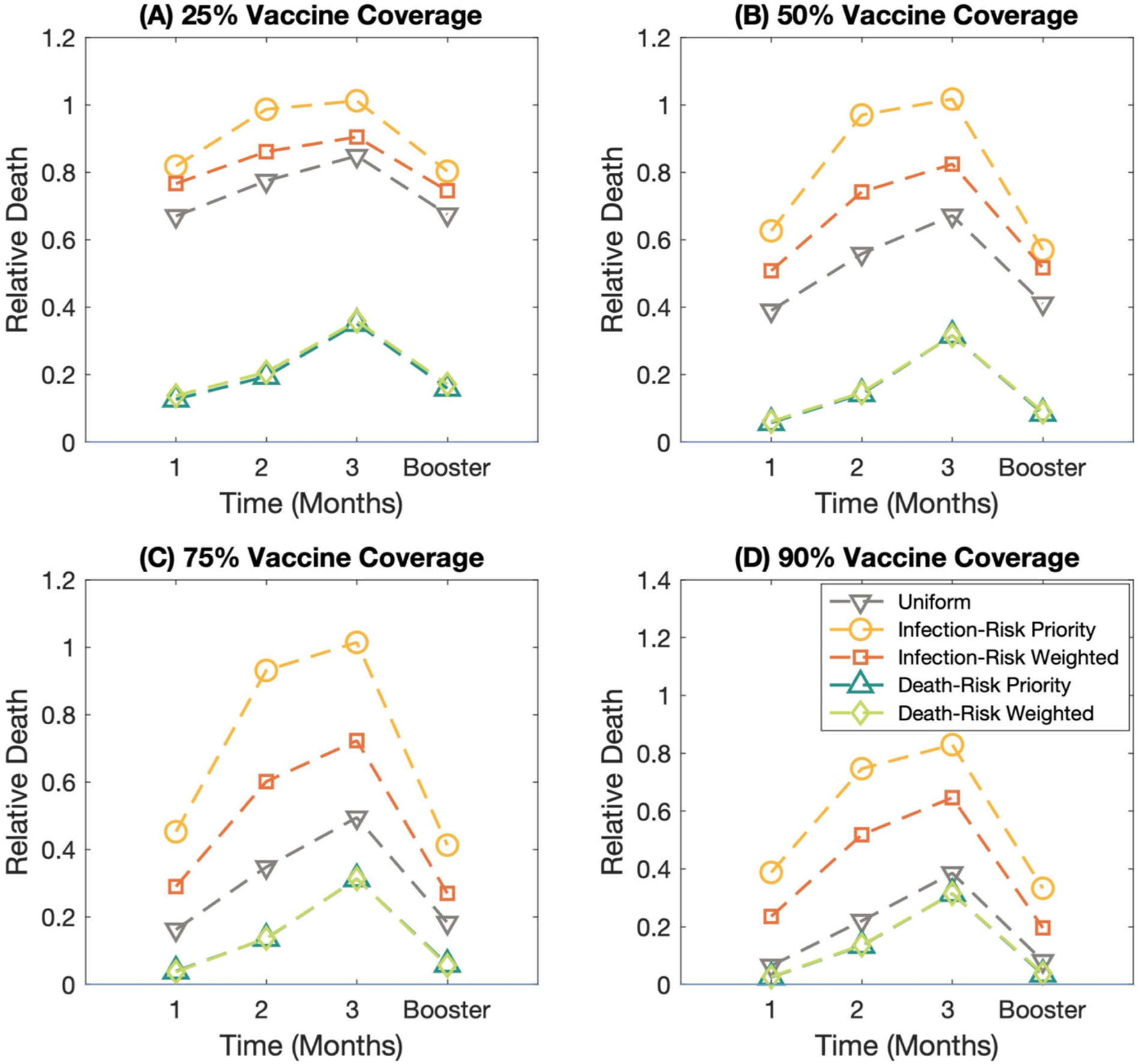
The effect of waning vaccine effectiveness against the Omicron variant (B.1.1.529) on the relative death. **(A) – (D)** display the extinction probabilities under the different vaccine coverages; 25%, 50%, 75%, and 90% of the total population, respectively. Each line represents the extinction probability of different vaccination strategies.

**Figures 6(A) – 6(D)** display the relative case at varied vaccine coverage: 25%, 50%, 75%, and 90%, respectively. We defined the relative cases as the cumulative cases at the extinction time compared to the cumulative cases at 0% vaccine coverage. The results display that the infection-risk priority vaccination strategies play a vital role in mitigating the cumulative cases in the first month after getting the full vaccine doses and after getting the booster shot. When the vaccine-induced immunity gets waned about 2-3 months after getting the full protection, the different vaccine allocation plans do not influence the relative cases, even if the vaccine coverage becomes greater.

**Figures 7(A)** – **7(D)** illustrate the relative death at 25%, 50%, 75%, and 90% vaccine coverage, respectively. We found that the waning vaccine effectiveness impacts relative death. Furthermore, vaccine allocation plans significantly affect the number of deaths, even though the vaccine effectiveness against the Omicron variant wanes. From **Figure 7**, the mortality-risk priority and the mortality-risk weighted vaccination strategies show great performance in reducing the number of deaths. The results also show that the higher vaccine coverage could mitigate the number of deaths, though the vaccine effectiveness gets waned.

## 4. Discussion

In this study, we investigated the impact of various vaccination strategies on the probability of extinction, cumulative cases, and deaths. We examined the robustness of the vaccination plan by varying the contact matrices and the population structures. We, besides, studied the effect of the waning vaccine effectiveness on the extinction probability, the cases, and the deaths.

We validated the method for investigating the extinction probability by comparing the results from the simulations to the one calculated from the theory under the condition that the vaccine effectiveness is perfect and the contact patterns in a population are random and uniform. The results confirm that the simulated results agree well with the theoretical ones. Then, we employed this method to examine the extinction probability in the heterogeneous system because the contact patterns are heterogeneous in the real world. The results indicate that heterogeneity plays a crucial role in driving the outbreak to go extinct with a higher chance. The heterogeneity indeed enhances the probability of outbreak extinction. It is consistent with the finding by Hagenaars T.J. *et al*. (2004) that increasing the level of heterogeneity results in decreasing in disease persistence [41]. This could be resulted from the difference between the contact rates among age groups. A lack of consistency causes the various transmission rates between multiple age groups. If an index case is in an elderly group, the chance of outbreak emergence is low, as the population in these groups has less social contact than those in working-age groups. On the other hand, the probability of outbreak emergence is high if an index case is in a working-age group because of more social contacts, not only in their age group but also through other age groups; hence, escalating the transmission rates.

To examine the consequence of various vaccination strategies on the extinction probability, the number of cases, and the number of deaths, we proposed five vaccination strategies; the infection-risk priority, the infection-risk weighted, the mortality-risk priority, and the mortality-risk weighted, and the uniform vaccination plans. The results reveal that the infection-risk priority vaccination strategy could maximize the extinction probability with the minimum vaccine doses required to prevent the emergence of disease outbreaks. Correspondingly, this strategy could remarkably reduce the cumulative cases. It can be visualized and explained by the concept of a contact matrix. When the population in the high infection risk age groups are immunized, the seed of the infectious disease appearing in other age groups has less ability to spread the disease. It reduces the transmission rate; consequently, it raises the chance of an outbreak going extinct. To relieve the rate of dying, both mortality-risk priority and mortality-risk weighted vaccine allocation strategies might be the optimal option. When the outbreak transmits rapidly, the vaccination strategies do not influence the extinction probability and the infection rate. However, the vaccine allocation strategies impact the reduction in deaths. Under the high vaccine effectiveness condition, the infection-risk priority vaccination plan still shows outstanding performance to maximize the extinction probability with the minimum vaccine doses and curb the cumulative cases. In addition, the infection-risk priority vaccination strategy could mitigate the deaths when the vaccine coverage is high. For low vaccine coverage, the mortality-risk priority and mortality-risk weighted strategies are effective vaccination plans to reduce the death rate.

To observe that our findings could be applied to multiple population structures and contact patterns, we deployed the population structures and contact matrices of low-, low-middle-, upper-middle-, and high-income countries in the determination. We examined whether the various vaccination strategy earns the same results under the different age-structured populations and contact patterns in the population. The results demonstrate that the infection-risk priority strategy is still a pleasurable vaccination plan to drive the outbreak to go extinct with the minimum number of vaccine doses and relieve the cumulative cases. In addition, the mortality-risk priority and the mortality-risk weighted strategies show good performance for alleviating the deaths. It indicates that the proposed vaccination strategies could be applied to distributing the vaccine in many nations. However, the results of uniform and infection-risk weighted vaccine allocation are not consistent for all population structures. It is due to the population structure. The infection-risk weighted vaccination strategy shows a better performance for reducing the number of deaths than the uniform vaccination strategy under the expansive population structure scenario, as the elders of the expansive population structure are less than other age bands. All seniors could get vaccinated when the vaccine is weighted by the infection risk. Additionally, the elders take a high risk of dying from this disease. It is reasonable that the infection-risk weighted vaccination strategy shows a better performance in reducing the number of deaths. For the constructive and stationary population pyramids, the uniform vaccination strategy shows better results for relieving the number of deaths. For these types of population structures, the number of populations is few differences between each age group. Distributing vaccines weighted by the infection risk could earn the benefit of mitigating the number of cumulative cases, but the uniform vaccine distribution plan might show the advantages of reduction in the mortality rate.

After massive vaccination was implemented to control COVID-19 transmission, multiple studies indicated that there is a drop in vaccine effectiveness over time [19–22]. Furthermore, the new SARS-CoV-2 variants could gain the immune escape potential and evoke a new wave of infections globally. We, hence, investigated the effect of waning vaccine effectiveness on the extinction probability, the cases, and the deaths. The results represent that various vaccination strategies do not help drive the outbreak to go extinct and mitigate the infection rate when the vaccine effectiveness drops. However, vaccination strategies still influence the rate of dying significantly. We, additionally, found that the mortality-risk priority and the mortality-risk weighted vaccination strategies show good performance in reducing the number of deaths. The results also reveal that the higher vaccine coverage could remarkably relieve the risk of dying and the risk of infection and enhance the probability of extinction under the waned vaccine effectiveness scenario.

Some limitations might exist in this study. For the infection-risk priority and mortality-risk priority vaccination strategies, the vaccines are prioritized for all populations in the age group with high infection and mortality risk, respectively. In the real world, distributing vaccines to every individual in a specific group while the people in other groups get vaccinated later might be impossible. Our model also included the age-specific contact patterns, but we did not consider human mobility or migration affecting the disease transmission [42–44]. In addition, we did not incorporate other control measures which might affect the infections [45–47].

## 5. Conclusions

In summary, our modeling results indicated that vaccination strategies affect the probability of outbreak extinction and the reduction in the number of cases and deaths. We found that the infection-risk priority vaccination strategy could enhance the extinction probability and mitigate the cumulative cases. To reduce the number of deaths, both mortality-risk priority and mortality-risk weighted vaccination strategies provide better results. However, the infection-risk priority might be a suitable strategy for constraining disease transmission and alleviating both cases and deaths under the high vaccine effectiveness scenario.

## Data Availability

All data produced in the present work are contained in the manuscript

## Author contributions

CM and NCJ designed the research and drafted the manuscript. CM, SC, and NCJ analysed the results and edited the manuscript. NCJ performed the simulations.

## Acknowledgments

We would like to gratefully acknowledge the valuable discussions with our colleagues and the members of the Biophysics Group, Department of Physics, Faculty of Science, Mahidol University. NCJ acknowledges the Development and Promotion of Science and Technology Talents Project for the scholarship support.

## Data Availability Statement

The data supporting the findings can be found in the main paper.

## Supplementary Information

### Disease transmission model

The rate of change of population in each compartment is governed by the following ordinary differential equations:

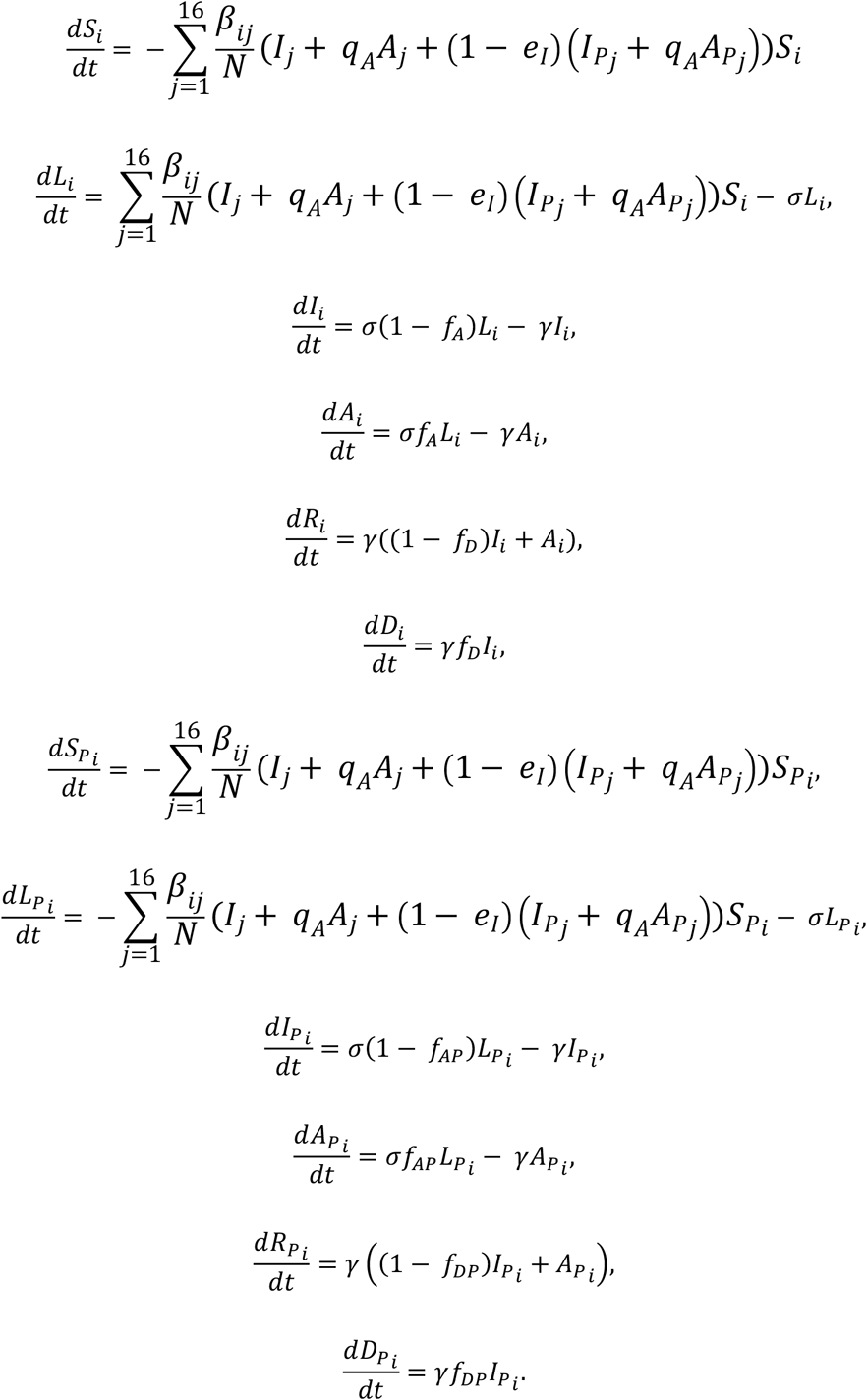

### Estimating the fraction of vaccinated asymptomatic infection

According to the definition of the vaccine efficacy against disease,

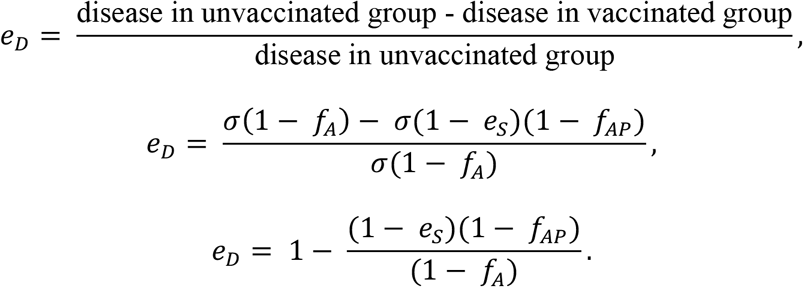

Therefore,

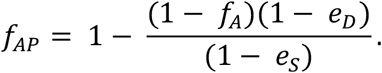

### Estimating the fraction of unvaccinated deaths

The infection fatality ratio (IFR) represents the proportion of deaths among all infected individuals, including all asymptomatic and undiagnosed infected individuals:

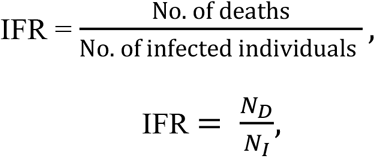

where *N_I_* = (1 − *f_A_*)*N_I_* + *f_A_N_I_* and *N_D_* = *f_D_*(1 − *f_A_*)*N_I_*

*f_A_* = Proportion of asymptomatic individuals

**f*_D_* = Proportion of deaths

*N_I_* = Number of infected individuals

*N_D_* = Number of deaths

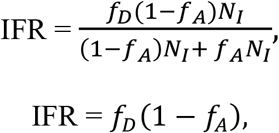

Thus,

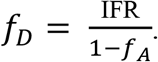

### Estimating the fraction of vaccinated deaths

Vaccine effectiveness is measured by calculating the risk of disease among vaccinated and unvaccinated persons and determining the percentage reduction in risk of disease among vaccinated persons relative to unvaccinated persons. The vaccine effectiveness against death is given by

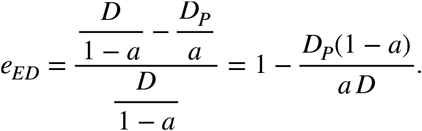

*e_DE_* = vaccine effectiveness against death

*D* = number of deaths in unvaccinated population

*D*_P_ = number of deaths in vaccination population

*f_A_* = Proportion of asymptomatic individuals

*f_AP_* = Proportion of asymptomatic vaccinated individuals

*f_D_* = Proportion of deaths

*e_S_* = Vaccine effectiveness against infection

*e_I_* = Vaccine effectiveness against transmission

*e_D_* = Vaccine effectiveness against symptomatic disease

*q_A_* = Asymptomatic Infectiousness

From the model,

*S* = 1 −*a*; *a* is the vaccine coverage (fraction of vaccinated population)

*S_P_* = (1 − *e_S_*)*a*

*S_V_* = *e_S_a*

Let

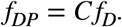

Then,

*D_P_* = (1 − *e_S_*)*aλσ1 0-* (1 − *f_AP_*)*γ f_D_C*;

*D* = (1 − *a*)*λσ* (1 − *f_A_*)*γ f_D_*

From

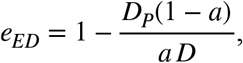

We got

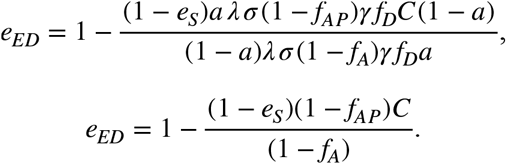

From

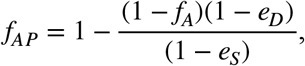

We rearranged it

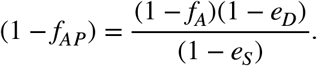

Thus,

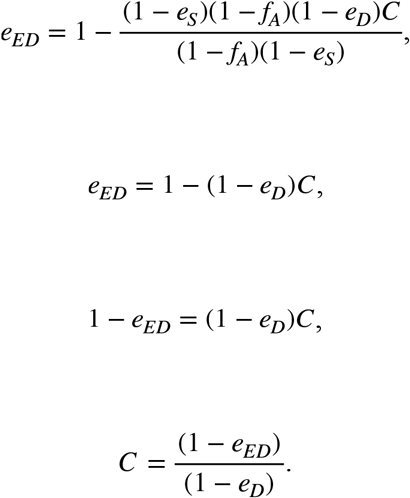

Finally, we got

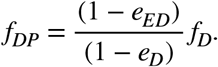

---

**Table S1:**
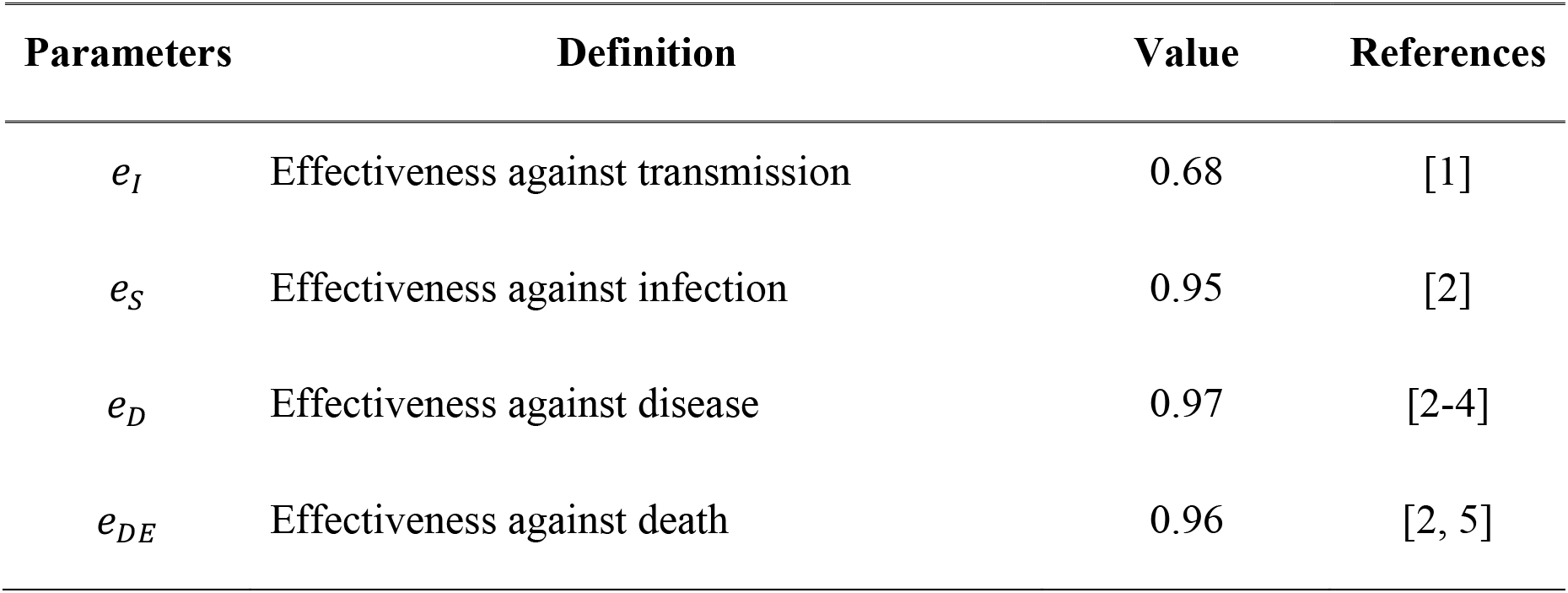
Parameters used for the high vaccine effectiveness scenario

| Parameters | Definition | Value | References |
| --- | --- | --- | --- |
| $e_I$ | Effectiveness against transmission | 0.68 | [1] |
| $e_S$ | Effectiveness against infection | 0.95 | [2] |
| $e_D$ | Effectiveness against disease | 0.97 | [2-4] |
| $e_{DE}$ | Effectiveness against death | 0.96 | [2, 5] |

**Figure S1:**
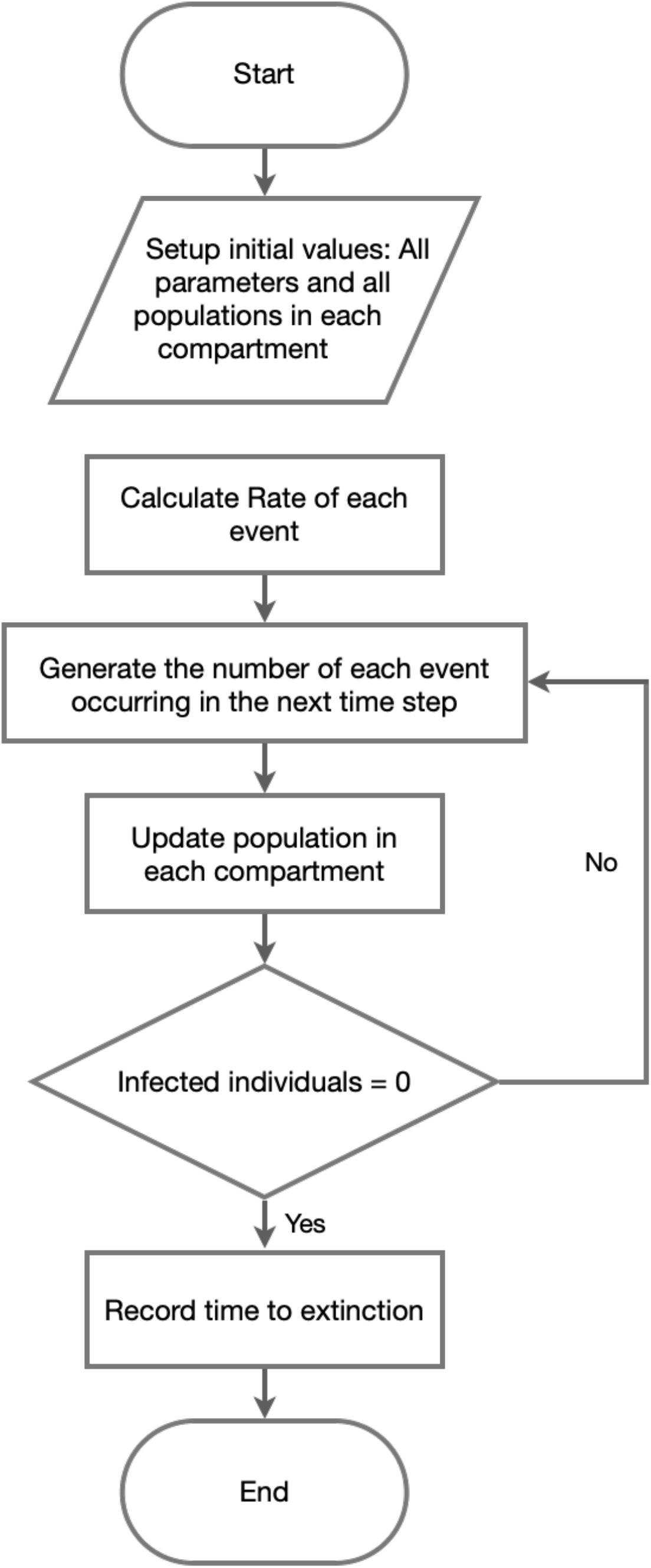
Model algorithm flowchart.

**Figure S2.**
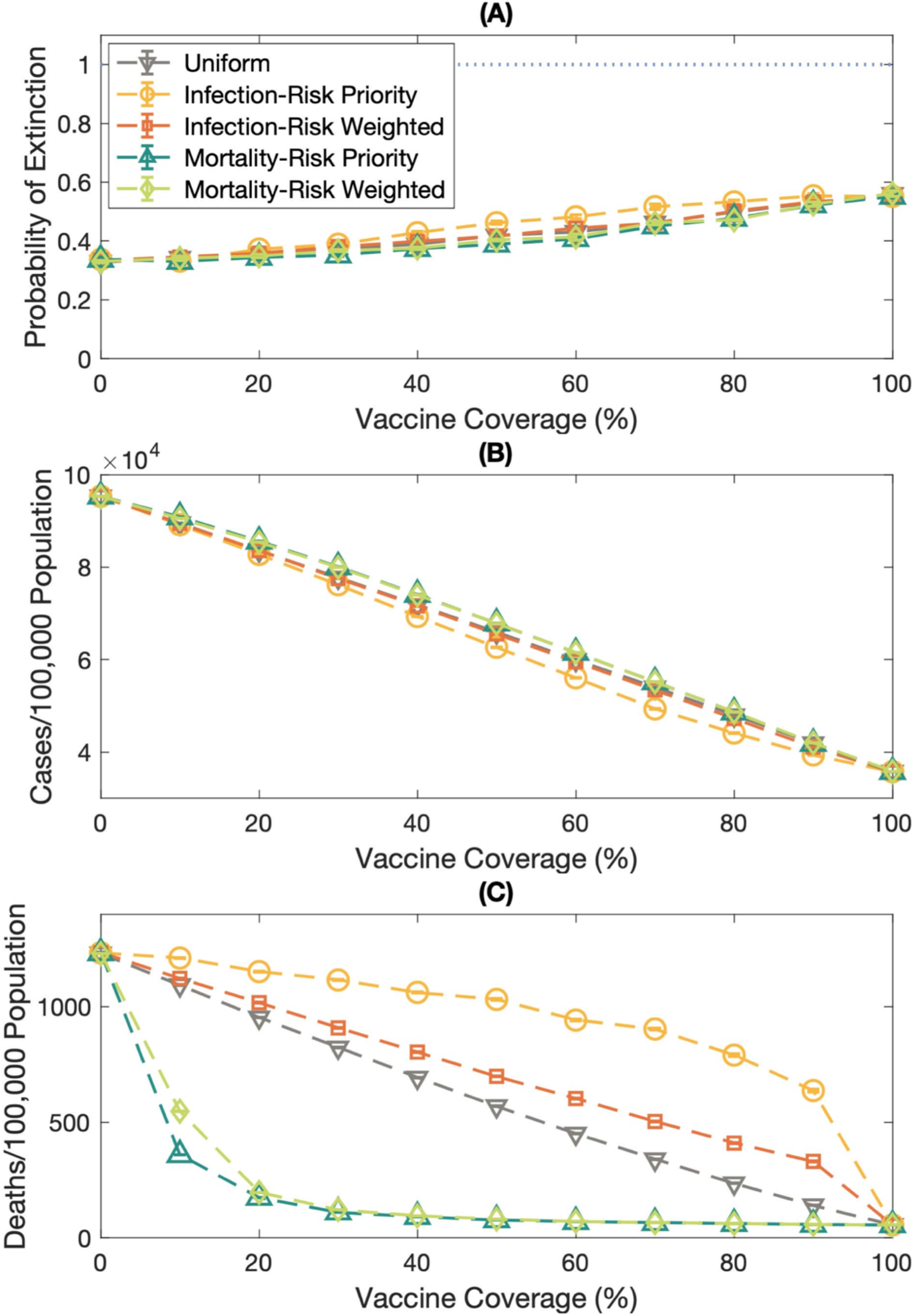
The impact of various vaccination strategies on the extinction probability, the infection rate, and the mortality rate under the high transmission rate, *R* = 5.0.

**Figure S3.**
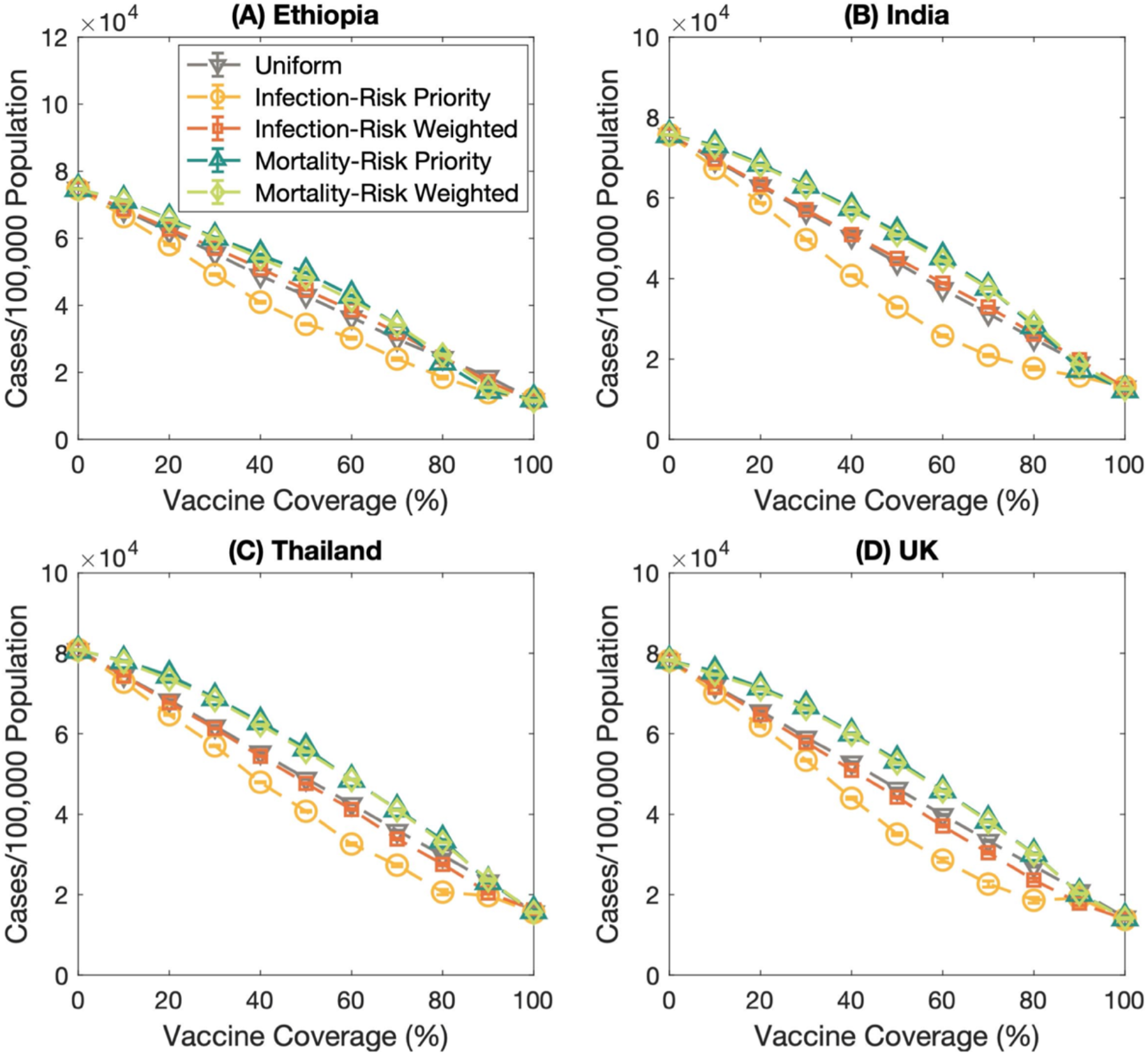
The number of cases per 100,000 population of various countries; **(A)** Ethiopia, **(B)** India, **(C)** Thailand, and **(D)** the United Kingdom. The dashed lines with the downward-pointing triangle, the circle, the square, and the upward-pointing triangle, and the diamond marks represent the number of cases per 100,000 population of the uniform, infection-risk priority, infection-risk weighted, mortality-risk priority, and mortality-risk weighted vaccination strategies, respectively. Error bars show the standard error.

**Table S2:** Vaccine effectiveness against Omicron variants. Data were obtained from [1, 6, 7].

| **month** | **es** | **e<sub>I</sub>** | **e<sub>D</sub>** | **e<sub>DE</sub>** |
| --- | --- | --- | --- | --- |
| 1 | 0.55 | 0.032 | 0.49 | 0.95 |
| 2 | 0.16 | 0.032 | 0.30 | 0.93 |
| 3 | 0.098 | 0.032 | 0.15 | 0.83 |
| **booster** |  |  |  |  |
| 1 | 0.55 | 0.077 | 0.67 | 0.95 |

